# The effect of remote ischemic preconditioning on postoperative cardiac and inflammatory biomarkers in pancreatic surgery: a randomized controlled trial

**DOI:** 10.1101/2020.12.18.20248465

**Authors:** Laura van Zeggeren, Remco A. Visser, Lisette M. Vernooij, Ineke M. Dijkstra, Madeleen Bosma, Izaak Q. Molenaar, Hjalmar C. van Santvoort, Peter G. Noordzij

## Abstract

**Background:** Cardiac and inflammatory biomarkers have been associated with adverse outcome after major abdominal surgery. Remote ischemic preconditioning (RIPC) may protect organs from ischemic insults during and after cardiac surgery, but the effect in major abdominal surgery is largely unknown.

**Objective:** To study the effect of RIPC on cardiac and inflammatory biomarkers in patients undergoing pancreatic resection.

**Methods:** Single-center, double-blind, randomized controlled trial in ninety patients undergoing elective pancreatic resection between March 2017 and February 2019. Three cycles of upper-limb ischemia and reperfusion (each 5 minutes) were applied before surgery. The primary endpoint was the maximum postoperative high-sensitive cardiac troponin (hs-cTn) T concentration within 48 hours after surgery. Secondary endpoints were postoperative myocardial injury (PMI, defined as a postoperative hs-cTnT ≥14 ng L^-1^), the maximum concentration of interleukin (IL)-6 within 48 hours after surgery, and postoperative complications within 30-days of surgery.

**Results:** RIPC did not reduce the maximum hs-cTnT concentration after surgery (12.6 ng L^-1^ vs 16.6 ng L^-1^ in the control group (P=0.23), nor did it lessen the incidence of PMI (15 (33.3%) patients in the RIPC group versus 19 (42.2%) controls, P=0.93). The maximum postoperative IL-6 concentration was 239 pg mL^-1^ [115-360] in the RIPC group, as compared to 317 pg mL^-1^ [174-909] in the control group (P=0.13). A postoperative complication occurred in 23 (51%) RIPC patients and 24 (53%) controls.

**Conclusions:** Remote ischemic preconditioning did not reduce the maximum postoperative hs-cTnT concentration. Postoperative myocardial injury, IL-6 concentrations and postoperative complications were not statistically different between RIPC patients and controls.

**Trial Registration:** Clinicaltrials.gov identifier NCT03460938

**Funding:** Funding for biomarker analysis was provided by Roche Diagnostics. Roche Diagnostics had no role in design and conduct of the study, analysis and interpretation of the data, preparation and approval of the manuscript.

**Article summary:** *Strengths and limitations of this study:*

1. Well-designed clinical trial in a selected group of high-risk abdominal surgery patients.
2. Serial assessment of high-sensitive cardiac troponin T and interleukin-6 concentrations.
3. Postoperative cardiac biomarker concentrations were relatively low.
4. This trial was not primarily designed to detect differences in IL-6 concentrations and postoperative complications.

## INTRODUCTION

Despite ongoing improvements in perioperative care, complication rates after major non-cardiac surgery remain substantial.^1-3^ The occurrence of a postoperative complication is an important determinant of poor functional recovery and long-term survival.^2, 4, 5^ Although the etiology of postoperative complications is not fully understood, myocardial injury has consistently been associated with poor outcomes after non-cardiac surgery, including major abdominal surgery, in a concentration-dependent manner.^1, 6^ An elevated cardiac troponin level, as a marker for postoperative myocardial injury (PMI), is often attributed to an oxygen supply demand mismatch (type 2 ischemia) in patients with coronary artery stenosis. However, the occurrence of PMI is inconsistent with the extent and severity of coronary artery disease.^7^ Preventive therapies aimed at reducing type 2 ischemia did not improve postoperative outcomes.^8, 9^

Besides ischemia, other pathophysiological perioperative mechanisms are related to PMI. Surgery can trigger a profound inflammatory response, which is characterized by high circulating levels of pro-inflammatory cytokines. Endothelium derived nitric oxide overproduction induces tissue hypoperfusion and mitochondrial dysfunction. The reperfusion injury that follows is an important cause of subsequent organ dysfunction.^10, 11^ High levels of inflammation in patients undergoing major abdominal surgery have previously been associated with PMI and postoperative complications.^12, 13^ However, no established treatments are available that protect organs from ischemia reperfusion injury due to inflammation during, or shortly after, non-cardiac surgery.

Ischemic preconditioning is a physiological mechanism that uses brief cycles of ischemia and reperfusion to protect organs from renewed ischemic insults. Studies in cardiac surgery patients have demonstrated a cardioprotective effect of remote ischemic preconditioning (RIPC) but were unable to show an improvement in postoperative outcome.^14-16^ This might have been the result of an interference with anesthetic drugs (i.e. propofol), although the exact mechanism of this interaction remains unclear.^17, 18^ The effect of RIPC on cardiac and inflammatory biomarkers in abdominal surgery is largely unknown. This study aimed to investigate the effects of RIPC on peak concentrations of high-sensitive cardiac Troponin T (hs-cTnT) and Interleukin (IL)-6 in patients undergoing pancreatic resection, as an example of major abdominal surgery.

## METHODS

### Trial design

The myocardial injury and complications after major abdominal surgery (MICOLON) 2 study was an investigator-initiated, single-center, randomized, double-blinded controlled trial. The study was conducted at St. Antonius Hospital, Nieuwegein of the Regional Academic Cancer Center Utrecht, a tertiary referral hospital for pancreatic surgery in the Netherlands with an annual volume of over 100 pancreatoduodenectomies. The study protocol was approved by the Medical research Ethics Committees United (MEC-U, number R16.042) and registered at clinicaltrials.gov (NCT03460938). The study was performed in accordance with the principles of the Declaration of Helsinki. All patients provided written informed consent. This study was performed and reported according to the CONSORT guidelines for randomized controlled trials.^19^

### Patients

Adult patients scheduled for elective pancreatic surgery (i.e. pancreatoduodenectomy, distal pancreatectomy and total pancreatectomy) between March 2017 and February 2019 were eligible for study participation. There were no exclusion criteria apart from pregnancy. All patients were screened in the outpatient preoperative anesthesia clinic by a physician or dedicated screening nurse. A full medical history and a physical examination was part of routine perioperative care in all patients.

### Trial procedures and blinding

Participants were randomly assigned on the day of surgery to the RIPC group or control group in a 1:1 ratio. Randomization was performed using a web-based system, in permuted blocks, with variable block sizes. RIPC was applied after induction of anesthesia and before incision by an appointed anesthesiologist, who was aware of the study-group assignment. Individual patients, surgeons and the investigators, who obtained and documented study data and clinical endpoints, were blinded to study group assignment. The RIPC protocol consisted of three 5-min cycles of upper-limb ischemia, which was induced by an automated cuff-inflator placed on the upper arm and inflated to 200 mmHg, with an intervening five minutes of reperfusion, during which the cuff was deflated. A sham procedure was performed in control patients, that consisted of a deflated cuff placed on the upper limb for 30 minutes.

### Perioperative care and blood sample analysis

Surgery was performed under general anesthesia and epidural analgesia, unless a contra-indication for epidural analgesia existed. To avoid interaction with ischemic preconditioning, propofol was not used for induction or maintenance of anesthesia. Instead, thiopental or midazolam were used for induction of anesthesia and sevoflurane for anesthesia maintenance. Further anesthetic management was left to the discretion of the attending anesthesiologist. All patients had an arterial line and central venous catheter for hemodynamic monitoring. Duration of surgery, fluid balance, blood loss, transfusion of blood products and use of inotropes or vasopressors were registered. After surgery, all patients were routinely admitted to an intensive care unit (ICU) until postoperative day (POD) 1. To improve recovery, patients were treated according to the Enhanced Recovery After Surgery (ERAS) guidelines.^20^ Discharge from the ICU was based on standard operating procedures and occurred at the discretion of the attending intensivist.

Blood samples were collected for the measurement of hs-cTnT and IL-6 concentrations after induction of anesthesia and before surgical incision (baseline) and at 4, 12, 24 and 48 hours. Blood samples were frozen and stored at −80°C until batch analysis. Analyses of both hs-cTnT and IL-6 were performed on an automated Cobas 8000 platform (Roche Diagnostics, Mannheim, Germany). Hs-cTnT analysis was done using a fifth generation Elecsys Troponin T high-sensitivity assay, IL-6 analysis was performed using the Elecsys IL-6 assay (Roche Diagnostics, Mannheim, Germany)

During the postoperative period, clinical data including postoperative complications were registered in the electronic patient record, as a part of standard care. Study data were entered in the RedCap [Vanderbilt University] database management system by investigators who were blinded for treatment allocation.

### Outcomes

The primary endpoint was the maximum postoperative hs-cTnT concentration within 48 hours after surgery. Secondary endpoints were the maximum postoperative concentration of IL-6 within 48 hours after surgery, PMI and postoperative complications within 30-days of surgery. PMI was defined as an absolute increase of hs-cTnT ≥14 ng L^-1^ above baseline concentration. A cut off value for an elevated IL-6 concentration was set at 432 pg mL^-1^ based on prior research.^12^ Postoperative complications were graded according to the standardized Clavien-Dindo (CD) classification of surgical complications and the Comprehensive Complication Index (CCI).^21, 22^ Pancreatic-specific complications (i.e. pancreatic fistula, bile leakage, postpancreatectomy haemorrhage, delayed gastric emptying, chyle leakage) were defined and classified according to the International Study Group of Pancreatic Surgery (ISGPS) definitions.^23-27^ Only grade B and C were reported as these are generally considered clinically relevant.

### Statistical analysis

Based on prior research from our institution on the association between postoperative hs-cTnT levels and complications in patients undergoing pancreatic resection (mean peak postoperative hs-cTnT 24 ng L^-1^ (±20)) we hypothesized that two groups of 45 patients were required to yield an 80% power and a significance level of 0.05, to demonstrate a 50% reduction in peak hs-cTnT concentration compared to the control group.^1^

Patients were analyzed based on the intention-to-treat principle. Baseline characteristics were described per treatment arm as percentages, means (± SDs), or medians (IQRs) as appropriate. Baseline differences between treatment arms were assessed and present for red blood cell (RBC) transfusion and duration of surgery. Multivariable analyses were used to adjust for this imbalance. For the primary outcome, linear regression was performed to compare mean peak postoperative hs-cTnT concentrations between both groups. Before analysis, hs-cTnT was log transformed and back-transformed geometric mean hs-cTnT concentrations were reported for both groups with its 95% confidence intervals (95% CI). Thereafter, we conducted generalized linear mixed models to analyze the effect of RIPC on postoperative hs-cTnT concentrations over time (i.e. at 4, 12, 24 and 48h after surgery). Univariate analyses were performed with time and treatment group as fixed parts and ‘subject’ as random part. To assess whether postoperative hs-cTnT measurements differed over time between both groups, an interaction term between treatment group and time was added to the fixed part of the model. Multivariable analysis was then conducted with adjustments for preoperative hs-cTnT, RBC transfusion and duration of surgery. Similar analyses were performed for the secondary outcome, i.e. postoperative IL-6 concentrations. As IL-6 trajectories were not linear over time, B-splines were applied using three knots for time to improve model fit. Models were compared based on the Akaike’s information criterion. Restricted maximum likelihood estimation was used to generate unbiased variance estimates for the final models.

Clinical outcomes, including postoperative complications, PMI and elevated IL-6, were compared using a chi square test, independent samples T test or Mann Whitney-U test, as appropriate. All analyses were performed according to the intention-to-treat principle. Two-sided P values of 0.05 or less were considered statistically significant. Data were analyzed using IBM SPPS software (version 24-26, for Windows) and R statistics (Version 3.5.1 – © 2018-07-02, R, Inc., for Windows).

## RESULTS

### Population

Ninety-two patients were randomized to the RIPC or control group. In two patients, surgery was cancelled due to peritoneal dissemination. Ninety patients were included in the final analysis, 45 patients in the RIPC group and 45 patients in the control group (Figure 1). Median age was 69 years and 59% of patients was male. A history of coronary artery disease (i.e. myocardial infarction or coronary revascularization) was present in 20% of patients (Table 1). The majority of patients had surgery for pancreatic cancer, with postoperative epidural analgesia. None of the patients were lost to follow-up and no RIPC related adverse events were observed.

**Table 1.** Baseline characteristics.

|  | RIPC<br>(N=45) | Control<br>(N=45) |
| --- | --- | --- |
| Median age (IQR) – yr | 69 (68-72) | 69 (65-73) |
| Male sex – no. (%) | 26 (58%) | 27 (60%) |
| Prior diagnosis – no (%) |  |  |
| Hypertension | 17 (38%) | 17 (38%) |
| Atrial fibrillation | 9 (20%) | 6 (13%) |
| Myocardial infarction | 8 (18%) | 3 (7%) |
| Coronary revascularization | 8 (18%) | 6 (13%) |
| Cardiac failure | 3 (7%) | 2 (4%) |
| Diabetes mellitus | 8 (18%) | 15 (33%) |
| Stroke | 1 (2%) | 4 (9%) |
| COPD | 5 (11%) | 4 (9%) |
| Renal insufficiency | 4 (9%) | 6 (13%) |
| Peripheral artery disease | 5 (11%) | 4 (9%) |
| ASA classification – no (%) |  |  |
| I | 1 (2%) | 5 (11%) |
| II | 23 (51%) | 23 (51%) |
| III | 19 (42%) | 15 (33%) |
| IV | 2 (4%) | 2 (4%) |
| Medications – no (%) |  |  |
| Sulfonylureas | 4 (9%) | 7 (16%) |
| Angiotensin receptor blockers | 6 (13%) | 4 (9%) |
| Nitrates | 3 (7%) | 3 (7%) |
| Aspirin | 10 (22%) | 6 (13%) |
| Type of surgery – no. (%) |  |  |
| Pancreatoduodenectomy | 35 (78%) | 39 (87%) |
| Distal pancreatectomy (± splenectomy) | 9 (20%) | 3 (7%) |
| Total pancreatectomy | 1 (2%) | 3 (7%) |
| Cancer surgery – no. (%) | 39 (87%) | 43 (96%) |
| Robot assisted surgery – no. (%) | 19 (42%) | 15 (33%) |
| Conversion to open procedure | 2 (4.7%) | 4 (9.1%) |
| Epidural catheter – no. (%) | 39 (87%) | 38 (84%) |
| Operating time – min. (mean, SD) | 329 (106) | 376 (110) |
| Blood loss – ml (IQR) | 400 (0 – 5100) | 400 (0 – 7000) |
| Intraoperative fluid balance – ml (IQR) | 2030 (150 – 11.000) | 2650 (-1500 – 8.000) |
| RBC transfusion – no. (%) | 3 (7%) | 10 (22%) |
| Use of inotropes and/or vasopressors – no. (%) | 42 (93%) | 39 (87%) |

ASA, American Association of Anesthesiologists
COPD, chronic obstructive pulmonary disease
PPPD, pancreaticoduodenectomy
RBC, red blood cell

**Figure 1.**
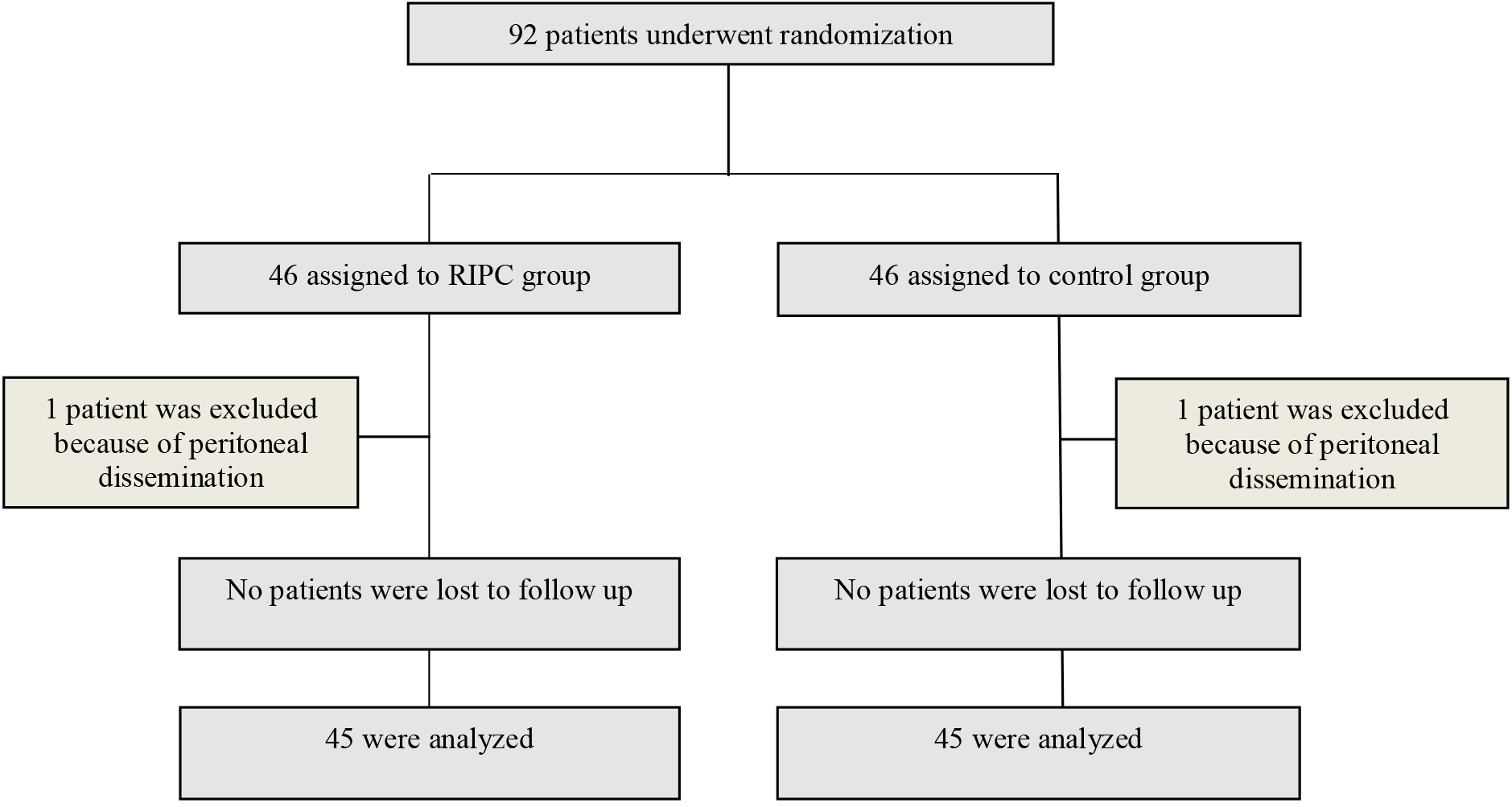
CONSORT flowchart. RIPC, remote ischemic preconditioning

### Cardiac biomarkers

Median preoperative hs-cTnT concentration was 9 ng L^-1^ [5-12 ng L^-1^] and not different between both groups (P=0.56). Before surgery, hs-cTnT was elevated (i.e. hs-cTnT ≥ 14 ng L^-1^) in 9 (20%) patients in the RIPC group, as compared to 10 (23%) patients in the control group (P=0.43). Remote ischemic preconditioning did not reduce the maximum hs-cTnT concentration after surgery (12.6 ng L^-1^ (95% confidence interval (CI) 5.9 – 27.0 ng L^-1^) versus 16.6 ng L^-1^ (95% CI 12.1 – 22.8 ng L^-1^)) in the control group (P=0.23), nor did it lessen the incidence of PMI (13 (28.9%) patients in the RIPC group versus 18 (40.0%) controls, P=0.375). Over time, postoperative hs-cTnT concentrations were not different between both groups (P=0.20, Figure 2). Perioperative hs-cTnT concentrations for individual patients are shown in Supplementary Figure A.

**Figure 2.**
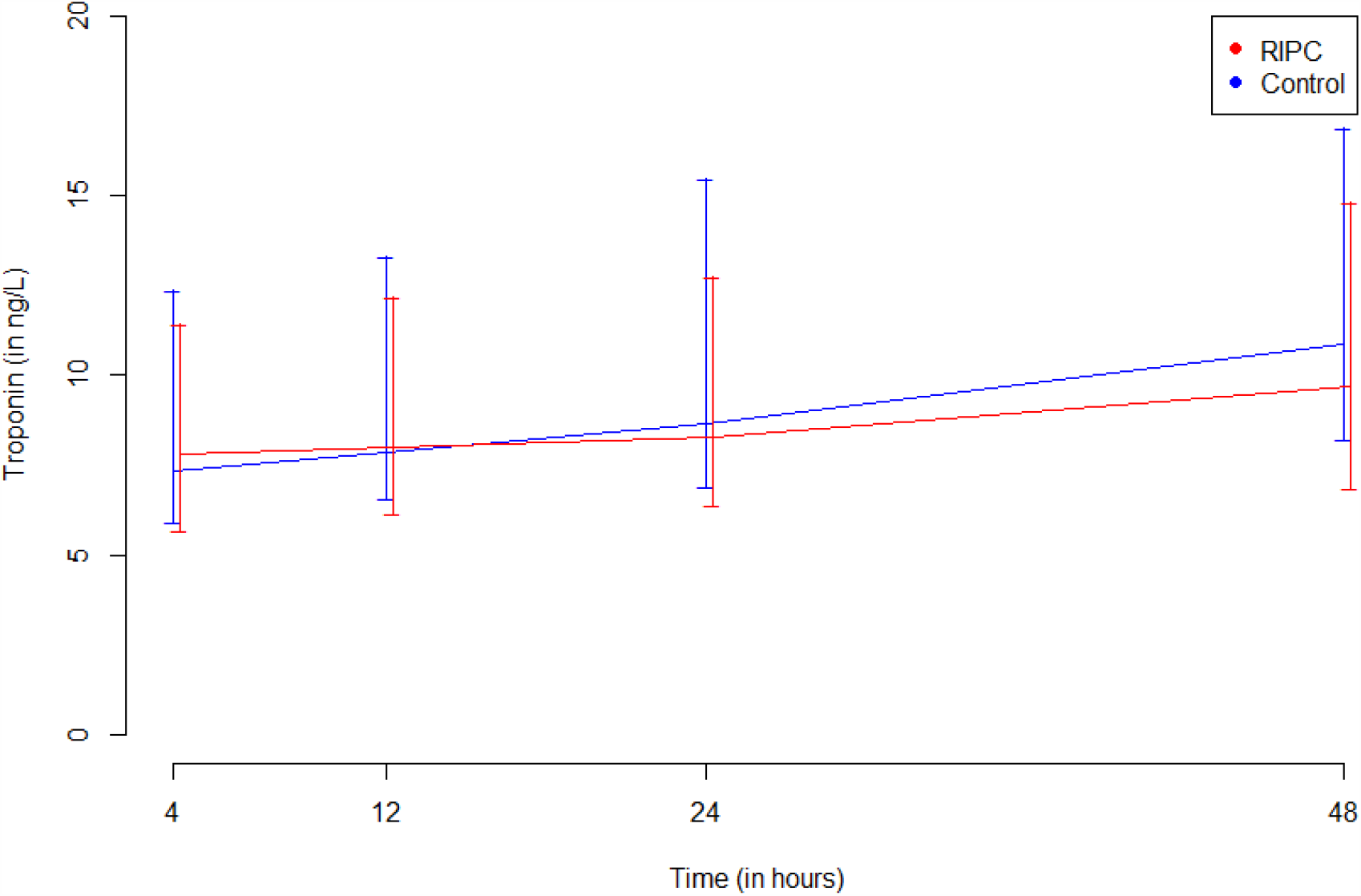
Postoperative high-sensitive cardiac troponin concentrations over time in the RIPC group and control group. Estimates are presented from the generalized linear mixed model, adjusted for preoperative hs-cTnT, red blood cell transfusion and duration of surgery. The 25^th^ and 75^th^ percentile are presented around each of the high-sensitive cardiac troponin measurements. Nine (2%) hs-TnT samples were missing. RIPC, remote ischemic preconditioning Hs, high-sensitive cTn, cardiac troponin

### Inflammatory biomarkers

Median preoperative IL-6 concentration was 4.4 pg mL^-1^ [3-7.7 pg mL^-1^] and did not differ between groups (P=0.93). The maximum postoperative IL-6 concentration was 239 pg mL^-1^ [115-360] in RIPC patients compared to 317 pg mL^-1^ [174-909] in controls (P=0.13). RIPC did not reduce the maximum absolute increase in IL-6 concentration, compared to baseline (265 pg mL^-1^ (95% CI: 122 – 1565 pg L^-1^)) vs 385 pg mL^-1^ (95% CI: 280 – 531 pg L^-1^), P=0.11). A postoperative IL-6 concentration >432 pg mL^-1^ was present in 13 (29%) RIPC patients and 16 (36%) controls (P=0.15). Over time, postoperative IL-6 concentrations were not different between the RIPC and control group (P=0.59, Figure 3). Perioperative IL-6 concentrations for individual patients are shown in Supplementary Figure B.

**Figure 3.**
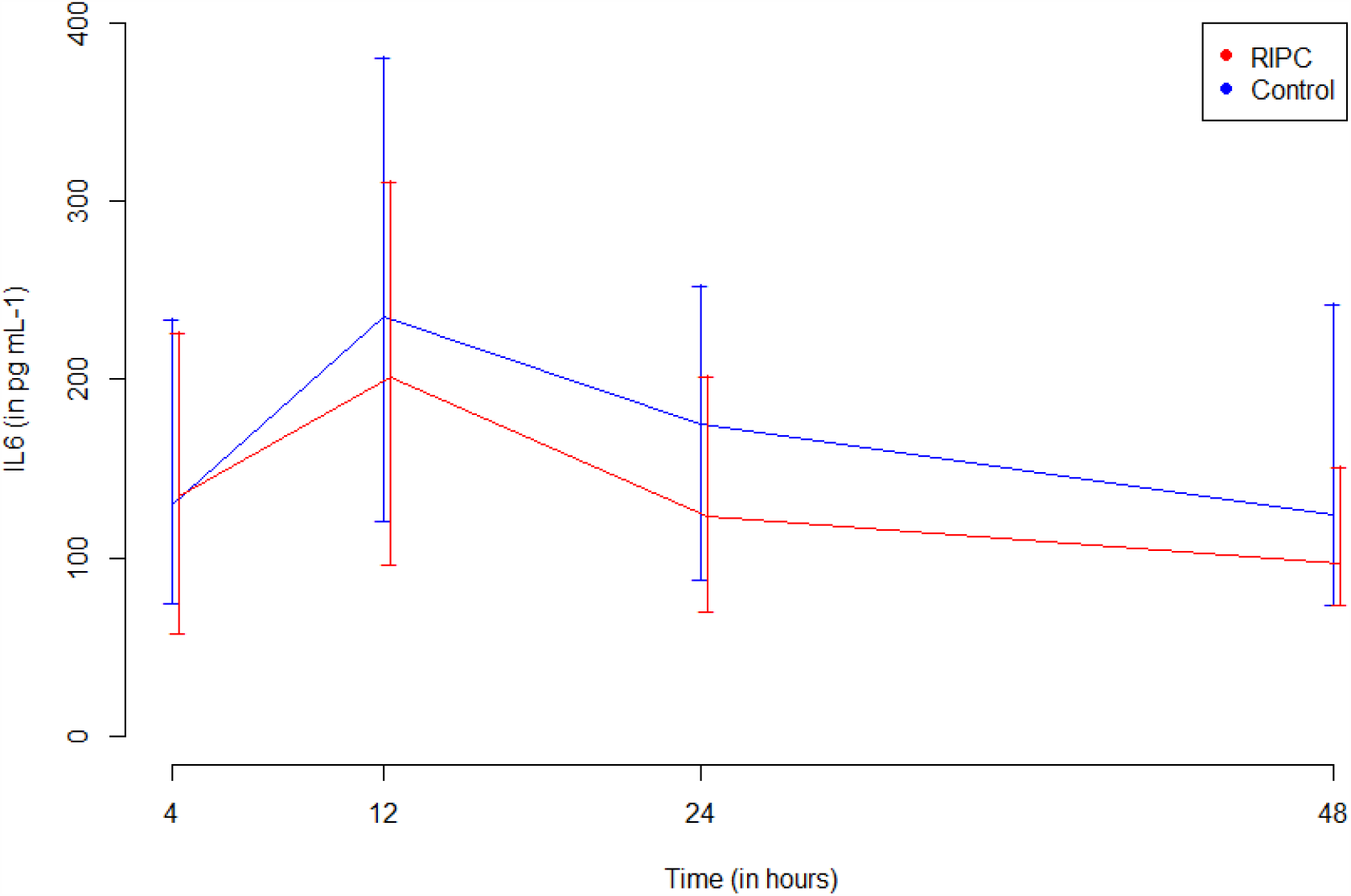
Postoperative interleukin-6 concentrations over time in patients in the RIPC group and control group. Estimates are presented from the generalized linear mixed model, adjusted for preoperative IL-6, red blood cell transfusion and duration of surgery. The 25^th^ and 75^th^ percentile are presented for each time interval. Nine (2%) IL-6 samples were missing. Red triangles represent mean IL-6 values. RIPC, remote ischemic preconditioning IL, interleukin

### Clinical outcomes

Median blood loss was 400 ml and 14% of patients received a red blood cell transfusion. The incidences of postoperative complications are presented in Table 2. A postoperative complication, defined as a CD class ≥ 3 or ISGPS class grade B or C, occurred in 23 (51%) RIPC patients, compared to 24 (53%) controls (NS). Median CCI was 27.6 [20.9-35.9] versus 30.8 [16.6-47.7] respectively (P=0.49). Length of ICU stay was 1.8 (±0.2) days in the RIPC group, as compared to 2.7 (±0.4) days in the control group; P=0.07). Overall length of hospital stay was 16.5 (±1.9) days versus 19 (±2.5) days respectively (P=0.35). Nine (20%) patients in the RIPC group were readmitted to the hospital within 30 days compared to 7 (16%) controls, P=0.58).

**Table 2.** Clinical outcomes.

|  | RIPC | Control | P value |
| --- | --- | --- | --- |
| Postoperative complication – no. (%) | (N=45) | (N=45) |  |
| Death | 0 | 1 (2%) | NS |
| Pancreatic fistula | 12 (27%) | 11 (24%) | NS |
| Bile leakage | 2 (4%) | 3 (7%) | NS |
| Post-pancreatectomy haemorrhage | 1 (2%) | 3 (7%) | 0.609 |
| Delayed gastric emptying | 10 (22%) | 9 (20%) | NS |
| Chyle leakage | 1 (2%) | 7 (16%) | 0.064 |
| Myocardial infarction – no. (%) | 0 | 0 | NS |
| Stroke – no. (%) | 1 (2%) | 1 (2%) | NS |
| Respiratory failure – no. (%) | 3 (7%) | 1 (2%) | 0.616 |
| Sepsis – no. (%) | 3 (7%) | 6 (13%) | 0.482 |
| Pneumonia – no. (%) | 0 | 1 (2%) | NS |
| Wound infection – no. (%) | 1 (2%) | 0 | NS |
| Urinary tract infection – no. (%) | 0 | 3 (7%) | 0.240 |

## DISCUSSION

In this double-blind randomized trial we studied the effect of RIPC on cardiac and inflammatory biomarkers in patients undergoing pancreatic surgery and showed that RIPC did not reduce the maximum postoperative hs-cTnT concentration. Furthermore, postoperative concentrations of IL-6 and postoperative complications were not statistically different between RIPC and control patients.

The cardioprotective effect of RIPC is well established in patients undergoing cardiac surgery. Recently performed large meta-analyses demonstrated that postoperative cardiac biomarker concentrations were lower in cardiac surgery patients after RIPC.^14-16^ To date, a cardioprotective effect of RIPC was not observed in patients undergoing major abdominal surgery. A previous randomized trial with RIPC in abdominal surgery patients (>90% of patients underwent colon surgery) found no differences in hs-cTnT concentrations until 72 hours after surgery.^28^ Although pancreatic surgery is considered a higher risk procedure than colon surgery (i.e. longer duration, more blood loss, higher rate of postoperative complications), and more patients had PMI in our study population (34% vs 21% in the previous study), we did not find a reduction in cTnT concentrations after RIPC. One could argue whether the degree of myocardial injury after abdominal surgery is large enough to show a protective effect of RIPC. As compared to cardiac surgery, peak cTn concentrations are 10 to 20 times lower than after major abdominal surgery. This may be partly explained by a relatively low prevalence of coronary artery disease in our cohort. Twenty percent of patients had established coronary disease (i.e. prior myocardial infarction or coronary revascularization) and a baseline hs-cTnT ≥ 14 ng L^-1^ was present in one out of five patients of the total study population. Also, minimal invasive robotic surgery, epidural anesthesia, and restricted transfusion management may have suppressed the ischemia reperfusion injury, which is targeted by RIPC. A possible interference of propofol with the cardioprotective effects of RIPC, which has been extensively described in prior studies, was ruled out by our study design. ^14, 16-18^

Surgical tissue injury triggers pro- and anti-inflammatory pathways. An excessive activation of the immune system, as reflected by high concentrations of IL-6, has been associated with adverse outcome after abdominal surgery.^12, 29^ Reports on the effect of RIPC on perioperative inflammation are scarce and primarily performed in cardiac patients. In 206 patients who underwent combined cardiac rhythm- and valve surgery, RIPC significantly decreased C-reactive protein (CRP) and neutrophil– lymphocyte ratio (NLR).^30^ Similar results were found in a cohort of 72 patients receiving radiofrequency ablation for atrial fibrillation, where the RIPC group showed a significant lower increase in CRP and IL-6.^31^ Other studies in patients undergoing major cardiac surgery, however, did not demonstrate an association between RIPC and postoperative concentrations of IL-6 and TNF-a.^32, 33^ The perioperative immune response is a complex process and is likely to differ largely between surgical procedures. Ischemia-reperfusion injury in pancreatic surgery may not be as extensive as in cardiac surgery, which was illustrated by the overall mild increase in perioperative IL-6 concentrations in our study population. This could explain the lack of effect of RIPC on the perioperative inflammatory response.

Our study has several limitations. First, postoperative concentrations of hs-cTnT were relatively low. This might be the result of the study design. Risk factors for coronary artery disease were not an inclusion criterion, in contrast to our previous study.^1^ As fixed coronary artery disease is believed to play an important role in PMI, this may have influenced our results. Second, our trial was not primarily designed (i.e. underpowered) to detect differences in IL-6 concentrations and postoperative complications. The lack of an effect on these outcomes may therefore be explained by a type two statistical error, since a non-significant attenuation in postoperative IL-6 concentrations and chyle leakage was indeed observed. Future research that focuses on the effect of RIPC on clinical outcomes and inflammation after pancreatic surgery may be worthwhile.

In conclusion, cardiac and inflammatory biomarkers after pancreatic surgery were not reduced by RIPC and no statistical difference was observed in clinical outcomes between both groups. This study adds information to the general discussion whether this theoretically promising, low cost, safe and easy to use technique, can be used to improve surgical outcomes.

## Data Availability

The principal investigators Laura van Zeggeren and Peter Noordzij had full access to all of the data in the study and take full responsibility for the integrity of the data and accuracy of the data analysis.

## ACKNOWLEDGEMENTS

Laura van Zeggeren: Data acquisition; Data analysis; Drafting the paper; Final approval

Remco A. Visser: Data acquisition; Final approval

Lisette M. Vernooij: Data analysis; Data interpretation; Critical revision; Final approval

Ineke M. Dijkstra: Critical revision; Final approval

Madeleen Bosma: Critical revision; Data acquisition; Final approval

Izaak Q. Molenaar: Critical revision; Final approval

Hjalmar C. van Santvoort: Critical revision; Final approval

Peter G. Noordzij: Study design; Data interpretation; Drafting the paper; Final approval

---

**Supplementary Figure A.**
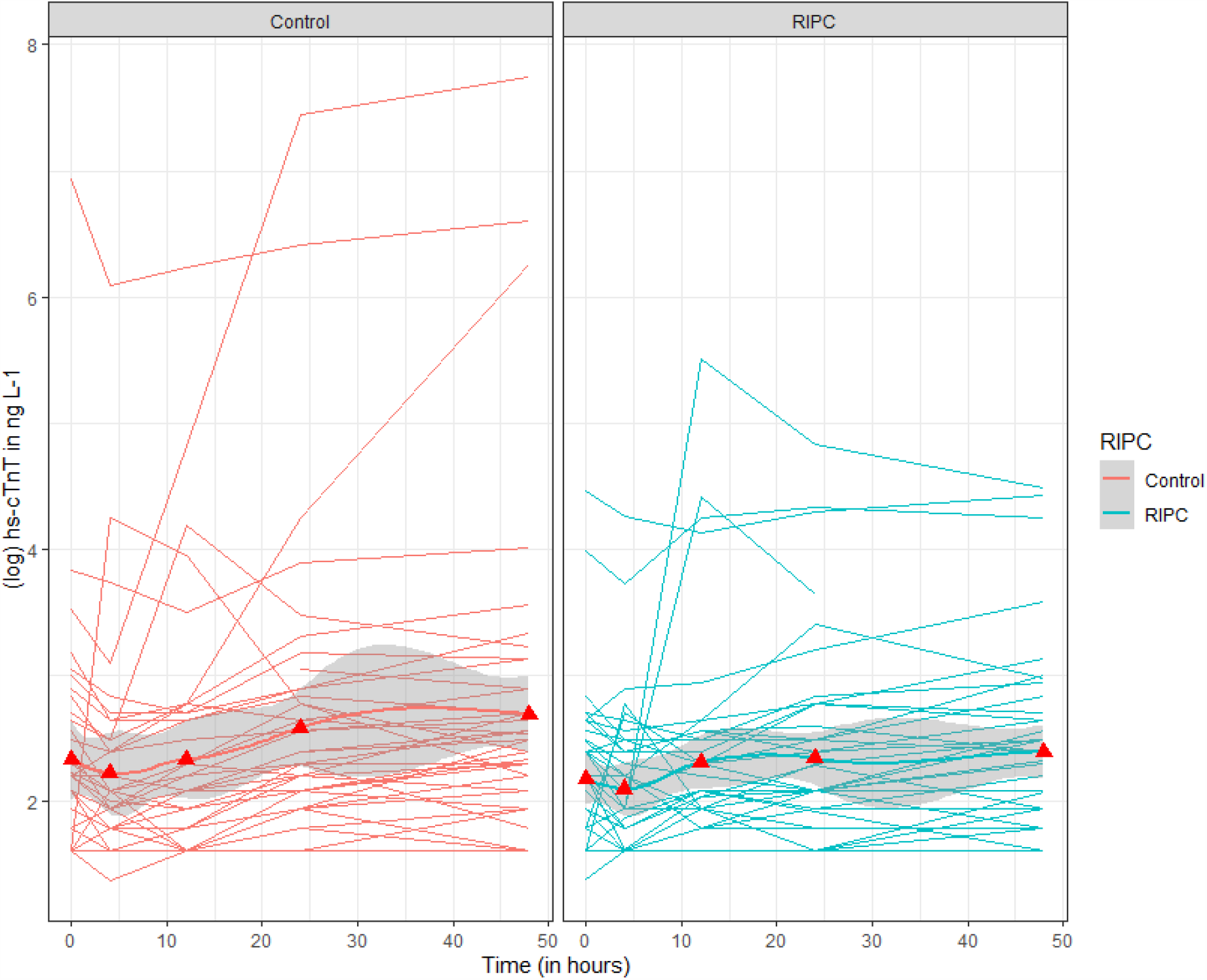
Observed high-sensitive cardiac troponin concentrations for individual patients in the RIPC group and control group. Red triangles represent mean high-sensitive cardiac troponin concentrations. RIPC, remote ischemic preconditioning Hs, high-sensitive cTn, cardiac troponin

**Supplementary Figure B.**
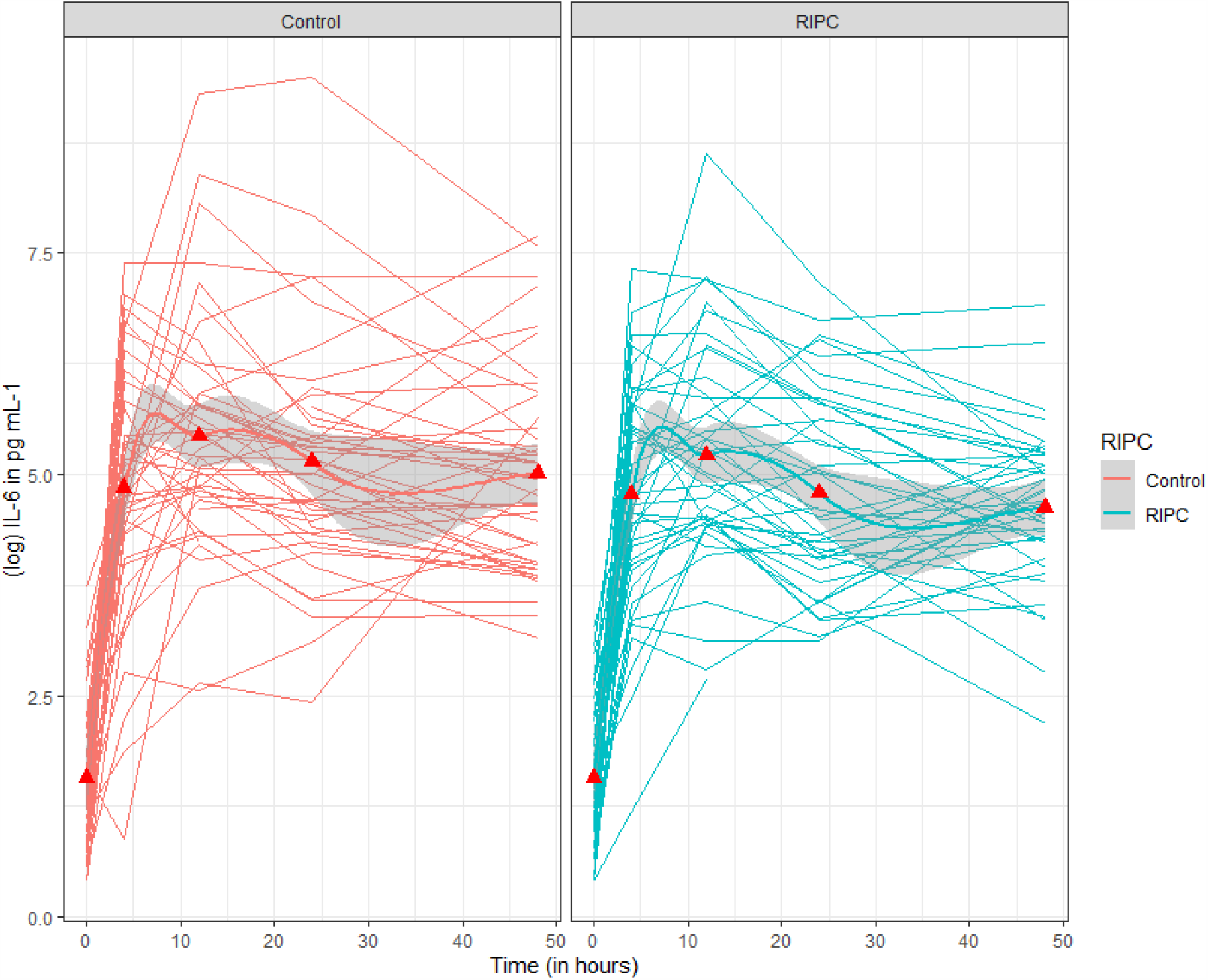
Observed interleukin-6 concentrations for individual patients in the RIPC group and control group. Red triangles represent mean interleukin-6 values. RIPC, remote ischemic preconditioning IL, interleukin

