## Supplemental figures for "The effect of remote ischemic preconditioning on postoperative cardiac and inflammatory biomarkers in pancreatic surgery: a randomized controlled trial"

**Supplementary Figure A. Observed high-sensitive cardiac troponin concentrations for individual patients in the RIPC group and control group**

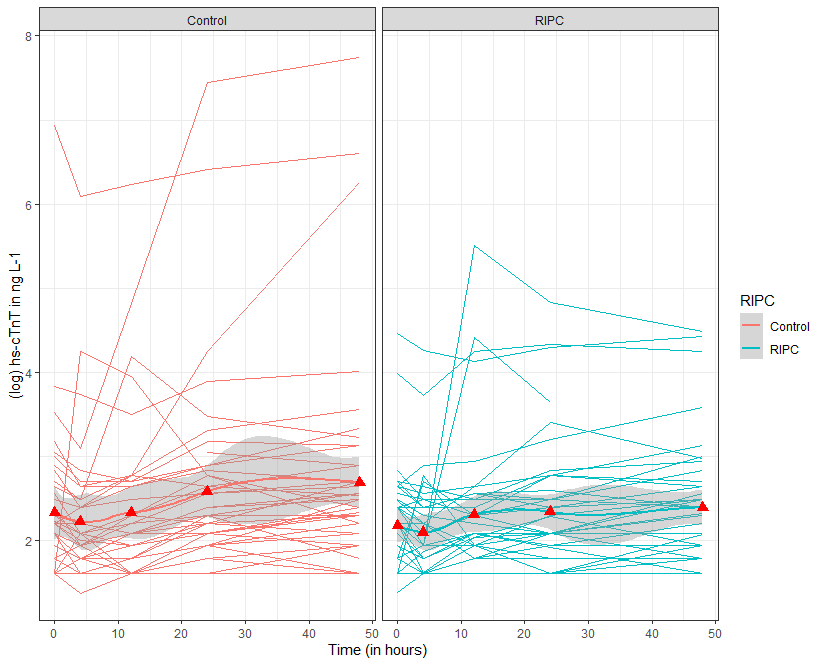

Red triangles represent mean high-sensitive cardiac troponin concentrations.

RIPC, remote ischemic preconditioning

Hs, high-sensitive

cTn, cardiac troponin

**Supplementary Figure B. Observed interleukin-6 concentrations for individual patients in the RIPC group and control group**

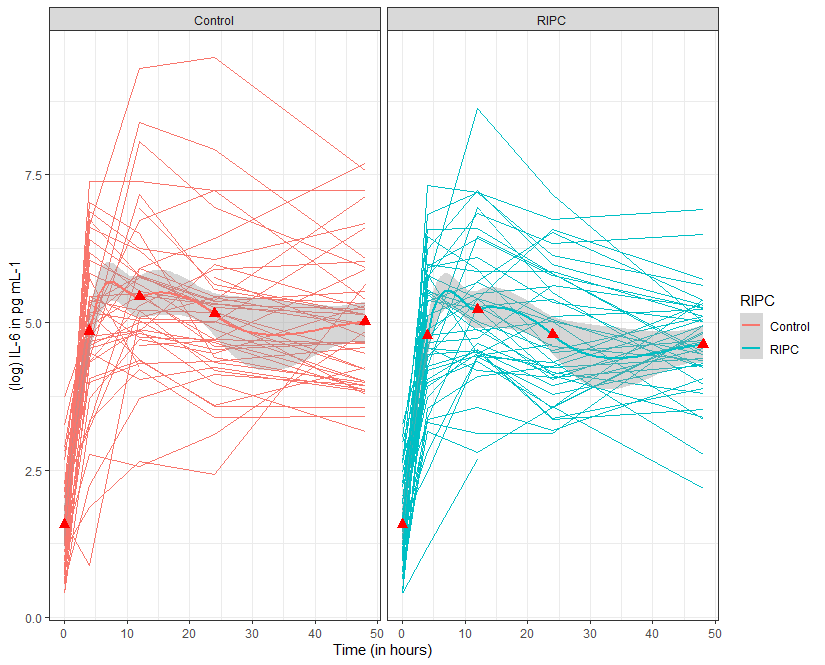

Red triangles represent mean interleukin-6 values.

RIPC, remote ischemic preconditioning

IL, interleukin
